# Prognostic Significance of CD8 T-cell Spatial Biomarkers in ER+ and ER-Breast Cancer

**DOI:** 10.1101/2025.05.27.25328389

**Authors:** Andrew E. Walker, Xiaohua Gao, Qichen Wang, Gabriela De la Cruz, Didong Li, Charles M. Perou, Joel Saltz, J.S. Marron, Katherine A. Hoadley, Melissa A. Troester

## Abstract

Tumor Infiltrating Lymphocytes (TILs) have been shown to be prognostic in Triple-Negative Breast Cancer (TNBC), but are rarely considered in other subtypes, particularly estrogen receptor (ER) positive cancers (70-80% of breast cancers), due to lower TIL counts. However, the spatial proximity of lower abundance TILs has not been evaluated in relation to breast cancer prognosis. In this study, multiplex immunofluorescent-stained images were used to identify cell types based on cytokeratin (Ck), CD8, and FoxP3 expression. Using distance-based visual morphometry between epithelial and immune cells, we computed new metrics, *proximity* and *consistency*, which capture spatial relationships between Ck+ tumor cells and CD8+ T-cells. Prognostic significance of proximity and consistency were compared to lymphocyte counts using log-rank tests. Worse relapse-free survival (RFS) was observed for both ER+ and ER-breast cancers with low proximity and consistency of CD8+ cells. Among ER negative breast cancers, proximity had the highest RFS hazard ratio (HR 1.84, 95% CI [1.18,2.87]). Among ER positive participants, RFS hazard ratios for proximity and consistency were 2.04 (95% CI [1.39, 2.98]) and 1.82 (95% CI [1.23, 2.69]) respectively. These associations were stronger than those observed for lymphocyte count (HR 1.35, 95% CI [0.92,1.98], log-rank p-value = 0.13). These IF-derived spatial metrics were also associated with established TILs metrics and RNA-based expression-based measures of tumor adaptive immune response. The prognostic significance of proximity in ER+ breast cancer implies that spatial parameters may identify individuals who would benefit from immune therapy; up to 75% of breast cancers experience T-cell proximity suggestive of immune susceptibility.

## Introduction

Studies over the past decade have suggested that the presence of tumor-infiltrating lymphocytes (TILs) is prognostically significant in breast cancer.^1–3^ In addition, immune therapy in conjunction with chemotherapy has been successful in treating Triple-Negative Breast Cancer (TNBC).^4,5^ However, 70-80% of breast cancers are ER+ and not eligible for immunotherapy under current guidelines.^6–8^ Nevertheless, these tumors often have moderate immune infiltration. Recently, several trials have demonstrated potential benefit of immune checkpoint inhibitors in high grade ER+/HER2-breast cancer.^9,10^ Given the ability to target the tumor immune microenvironment (TIME) and the potential benefits of immune therapy in combination with adjuvant chemotherapy, there is a need to better understand TIME across breast cancer subtypes.^5,11^

Our previous work has linked genomic instability with immune response in both ER+ and ER-participants, suggesting that immunogenic genomic instability is prognostically significant even in ER+ participants.^12^ However, immunogenic responses are difficult to detect in ER+ breast cancers as the TIL counts tend to be present at low or moderate levels. However, it may be important to consider whether immune cells are in close proximity to the tumor cells. In context of the high-count immune infiltration of ER-breast cancers, the assumption of proximity is more readily met. However, in moderately infiltrated tumors, distance between tumor and immune cells may vary. Very few studies have evaluated proximity or other spatial characteristics and few have used specific cell-type markers in analyses^11,13^, such as those available via multiplex immunofluorescent (IF).^14^ Reliance on TILs counts from H&E or bulk gene expression ^2,3,15^ ignore the potentially important spatial relationship between CD8+ cells and tumor cell death.^16^

We hypothesized that the distribution of lymphocytes with respect to tumor cells predicts prognosis. We considered proximity (measure of mean distance between CD8+ and Ck+ tumor cells) and consistency (variability in mean distances between CD8+ and Ck+ tumor cells) as two novel spatial metrics describing the distribution of CD8+ T-cells within the TIME. We compared these measures to established gene expression and count-based measures for breast cancer immune microenvironments and evaluated associations with breast cancer prognosis in a diverse study population of breast cancer patients.

## Methods

### Study Population

The data consists of participants from Phase 3 (2008-2013) of the Carolina Breast Cancer Study (CBCS). Methods of data collection for Phase 3 CBCS have been previously described in detail.^17^ Participants were recruited using rapid case ascertainment from the North Carolina Central Cancer Registry.^18^ CBCS eligibility required participants to be females aged 20-74 with their first diagnosis of invasive breast cancer and they must be residents of specified counties in North Carolina.^18^ Participants were enrolled under the University of North Carolina at Chapel Hill School of Medicine Institutional Review Board protocol and provided informed written consent.^18^ Multiplex IF images of TMA cores as well as time to recurrence data was available for 1740 participants. The dataset was reduced to 1467 participants after removing TMA cores which did not pass quality control standards, as described in the image processing section. Of the 1467 participants, 914 have Immune Class data^19^, and 864 have AGI/NGI classes^12^, and only 288 participants had WSI TIL spatial scores^20^. Data from all 1467 participants was used in the time to recurrence analysis, whereas only data from 914 participants was used when referencing Immune Classes, data from 864 participants was used when referencing AGI status, and only 288 participants were used when referencing the WSI TIL spatial scores. For the WSI data the 288 participants are ER positive by IHC expression and/or in the PAM50 Luminal B and HER2 molecular subtypes.^20^

### TMA Staining and Processing

Up to four 1mm cores were cut from a Formalin-fixed, paraffin-embedded (FFPE) tissue block and added to a Tissue Microarray (TMA) grid, and tissues were obtained from a subset of CBCS3 cases. Details of CBCS3 TMA construction have been previously described.^21^ Triple immunofluorescence (IF) was performed on 5 micron FFPE sections using the Bond Rx fully automated slide staining system (Leica Biosystems Inc.) and Bond Research Detection kit (DS9455). Slides were deparaffinized in Leica Bond Dewax solution (AR9222), hydrated in Bond Wash solution (AR9590) and sequentially stained for CD8 (Cell Marque, 108R-14), FoxP3 (abcam, ab20034), and CK (Leica Biosystems, NCL-L-AE1/AE3-601). Specifically, antigen retrieval for CD8 was performed for 20 min at 100° C in Bond-Epitope Retrieval solution 1 (ER1) pH 6.0 (AR9961). After pretreatment, slides were incubated for 1 hour with CD8 (1:200) followed with Novolink Polymer (Leica Biosystems, RE7260-CE) then TSA Cy3 (Akoya Biosciences, SAT704A001EA). A second round of antigen retrieval was performed for 10 min at 100° C in Bond-epitope retrieval solution 1 (ER1). Slides were then incubated with the FoxP3 (1:120, 2 hours) then Novolink Polymer or Post Primary plus Polymer combination and detected with TSA Cy5 (Akoya Biosciences, SAT705A001EA). A final round of antigen retrieval was performed with ER1 for 10 min before incubating the slides in CK (1:300, 30 minutes) followed by Novolink Polymer or Post Primary plus Polymer combination and Alexa Fluor™ 488 Tyramide Reagent (Thermo Fisher Scientific, B40953). Nuclei were stained with Hoechst 33258 (Invitrogen). The stained slides were mounted with ProLong Gold antifade reagent (Thermo Fisher Scientific, P36930). Positive and negative controls (no primary antibody) were included in this run. High resolution acquisition of IF slides was performed with the Aperio Versa 200 scanner (Leica Biosystems Inc.) at an apparent magnification of 20X. Images were uploaded to the eSlideManager database (Aperio). The staining process was done by Pathology Services Core at University of North Carolina at Chapel Hill (UNC), and this study was approved by the UNC Office of Human Ethics and Institutional Review Board.

### Image Processing

Multiplex IF images consist of blocks of 1mm TMA cores, where most participants have up to four cores and a small group of participants, who had two tissue samples taken, have up to eight cores. The images have four color channels with each color indicating one of four biomarkers: Hoechst (blue), Cytokeratin (cyan), CD8 (green), and FoxP3 (red). The Hoechst channel is an indicator for the cell nucleus, CD8 and FoxP3 are indicators for lymphocytes, and Cytokeratin is an indicator for tumor cells.

Cell segmentation and classification was performed in Qupath digital pathology software.^22^ Cells were segmented using the Cell detection function in Qupath which segments the cells based on the intensity of the Hoechst channel. Pathologists visually assessed several samples to confirm that the final segmentation gave a reasonable representation. Cell classification was performed using the simple threshold classifier in Qupath. The cells are classified as tumor, CD8 T cell (CD8), T regulatory cell (Treg), or unclassified. Thresholds were chosen based on careful analysis of the pixel level average channel intensity within the cell boundary. The cell is classified as Treg when the average intensity for both the CD8 and FoxP3 channel exceeded the threshold, CD8 when the average intensity of the CD8, but not the FoxP3, exceeded the threshold, and tumor only when the average intensity for the CK channel was above the threshold. A cell cannot be both a tumor cell and a lymphocyte, therefore any cell which exceeded both the threshold for CK and the threshold for CD8 or FoxP3 is classified as a lymphocyte.

Pathologists visually assessed several sample images to validate the final classification. Details on parameters used in the cell detection function as well as the cell classification thresholds that are used are available in the supplemental material (Supplemental Table 2). The centroid coordinates of the cells as well as the class labels were exported from Qupath and the proximity and consistency biomarkers were computed in Python as discussed in the next section. To ensure the quality of the data, any TMA core with fewer than 1000 tumor cells and fewer than 3000 total cells were removed.

### Spatial Biomarkers

The spatial biomarker values were computed in Python. The Nearest Neighbor Distance (NND) from a tumor cell to the CD8 T cell was computed for each TMA core. All distances were less than 1000 microns, based on the size of the TMA cores. Due to the highly skewed distribution of the NNDs, the base 10 logarithm (log10) of NND was determined to be a better representation. The collection of log10 NNDs for each TMA core corresponding to a single study ID were aggregated and the mean and variance for the distribution of aggregated log10 NNDs was computed for each study ID. The mean and variance of log10 NNDs are not intuitive measurements of proximity to tumor cells and consistency of distances. Instead, proximity is computed as a function of the mean of log10 NNDs and consistency is computed as a function of the variance of log10 NNDs to ensure, for example, that high values of proximity imply on average CD8 T cells are close to tumor cells and high values of consistency imply less variability in the distances between CD8 T cells and tumor cells. Proximity is given by the formula: (3 – mean of log10 NNDs from tumor cell to CD8 T cell). Proximity can range from 0 to 3 since the maximum mean of log10 NNDs is less than 3. Consistency is given by the formula: (1 – variance of log10 NNDs from tumor cell to CD8 T cell). Consistency ranges from 0 to 1 since the maximum variance of NNDs is less than 1. The lymphocyte count is computed as the total number of cells which were classified as CD8 T cells and Treg cells. When participants are split based on high/low lymphocyte count, the median count is used as the cutpoint. Proximity and consistency are binarized for the purpose of comparing differences in recurrence between participants with high vs. low values of proximity/consistency. We explored various cutpoints for proximity and consistency. The cutpoint used in the analysis was based on splitting the data into 4 groups based on the quartiles and then collapsing groups which had similar hazard ratios. After collapsing groups, the median was the final cutpoint for consistency and the 1^st^ quartile was the final cutpoint for proximity. An attempt was made to find an optimal cutpoint using a training and testing split, but this led to unstable results due to a only a small fraction of participants having a recurrence. Supplementary Table 3 provides the results from the attempted optimization.

### Statistical Analysis

The Kruskal-Wallis Test, a nonparametric version of the ANOVA test, was used to determine whether there are statistically significant differences between the medians of the proximity, consistency, and log10 lymphocyte count distributions by ER status as well as among breast cancer subtypes. The Kaplan-Meier method was used to estimate the RFS curves. All curves include estimates of 95% confidence bounds.

The log-rank test was used to compute the p-values for difference between survival curves. The hazard ratios and corresponding 95% confidence intervals are computed using the Cox proportional hazards model. In all cases, the proportional hazards assumption was confirmed using the proportionality test described by Grambsch and Therneau (1994)^23^ as implemented by the cox.zph function in the R Survival package. The likelihood ratio test was used for determining significance when comparing models in the case where the set of covariates of one model is a subset of the more complex model. Spearman’s correlation coefficient was used to determine monotonic association between our count, proximity, and consistency metrics and the Intratumoral Strength, Peritumoral Strength, TIL forest, and TIL desert scores. Conditional odds ratios and 95% confidence intervals were computed to determine the level of association of proximity, consistency, and lymphocyte count with AGI classified participants and to determine the level of association of proximity, consistency, and lymphocyte count with Adaptive-Enriched participants. The 0.05-level is used for all tests of statistical significance.

## Results

### Proximity and Consistency

To quantify lymphocyte infiltration, we computed the distances from each tumor cell to the nearest CD8+ lymphocyte (Figure 1). CD8+ cells were defined as positive for the CD8 marker and negative for FoxP3, which indicates a T-regulatory cell. Proximity was computed as an affine function of the mean of the log10 distances and consistency was an affine function of the variance of the log10 distances. All distances were computed on 1mm TMA cores with a majority of distances under 100 microns.

**Figure 1:**
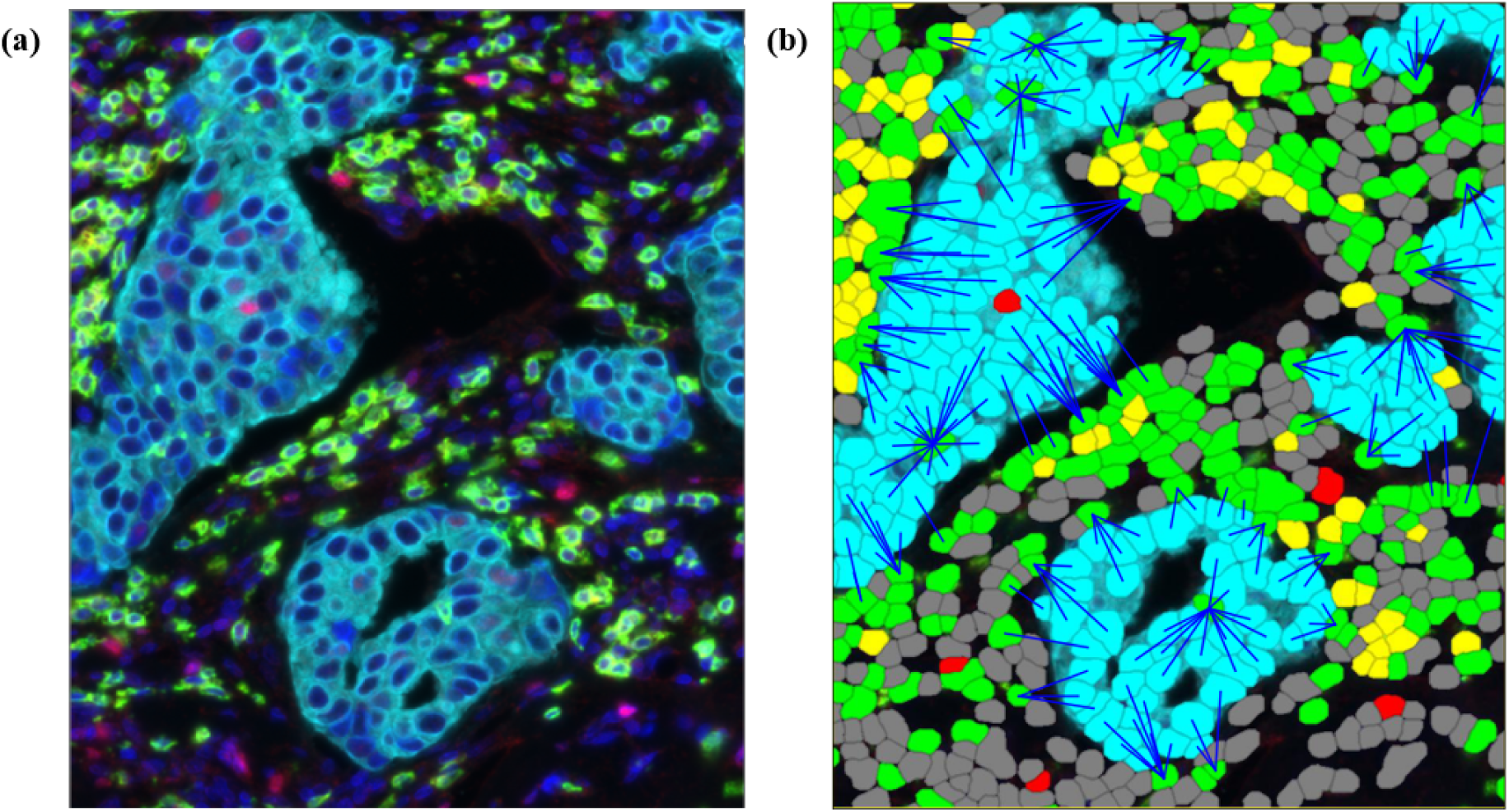
Multiplex IF image of breast cancer tissue. (a) Cells are stained with 4 colors indicating biomarkers for Hoechst (blue), Cytokeratin (cyan), CD8 (green), FoxP3 (red). (b) Multiplex IF image with overlayed colored tiles indicating cell classification: Epithelial (cyan), CD8 T cell (green), Treg (yellow), FoxP3 only (red), and unclassified (dark gray). Cells are detected using the Hoechst channel and classified as epithelial, lymphocytes, or unclassified based on the intensity of the Cytokeratin, CD8, and FoxP3 channels. Distance is computed from each epithelial cell to the nearest CD8 lymphocyte (length of blue line). The distances are used to compute proximity and consistency.

Lymphocyte count, defined as number of CD8 or FoxP3 positive cells, was computed for comparison to proximity and consistency since count is a commonly used spatial metric.

We computed kernel density estimates of the distributions of proximity, consistency, and count (Figure 2). The distributions of proximity and count tended to be wider while consistency had a much tighter distribution. The H-statistic given by the Kruskal-Wallis test was used to determine if there were statistically significant differences between the median of the count, proximity, and consistency by ER status (Figure 2). High proximity, high consistency, and high count were significantly associated with ER-negative status. Though there were statistically significant differences for all spatial markers by ER status, the log10 lymphocyte count had the strongest association with ER status (H = 76.44, p-value = 2.3e-18) followed by proximity (H = 17.45, p-value = 3e-5) and consistency (H = 6.01, p-value = 0.01).

**Figure 2:**
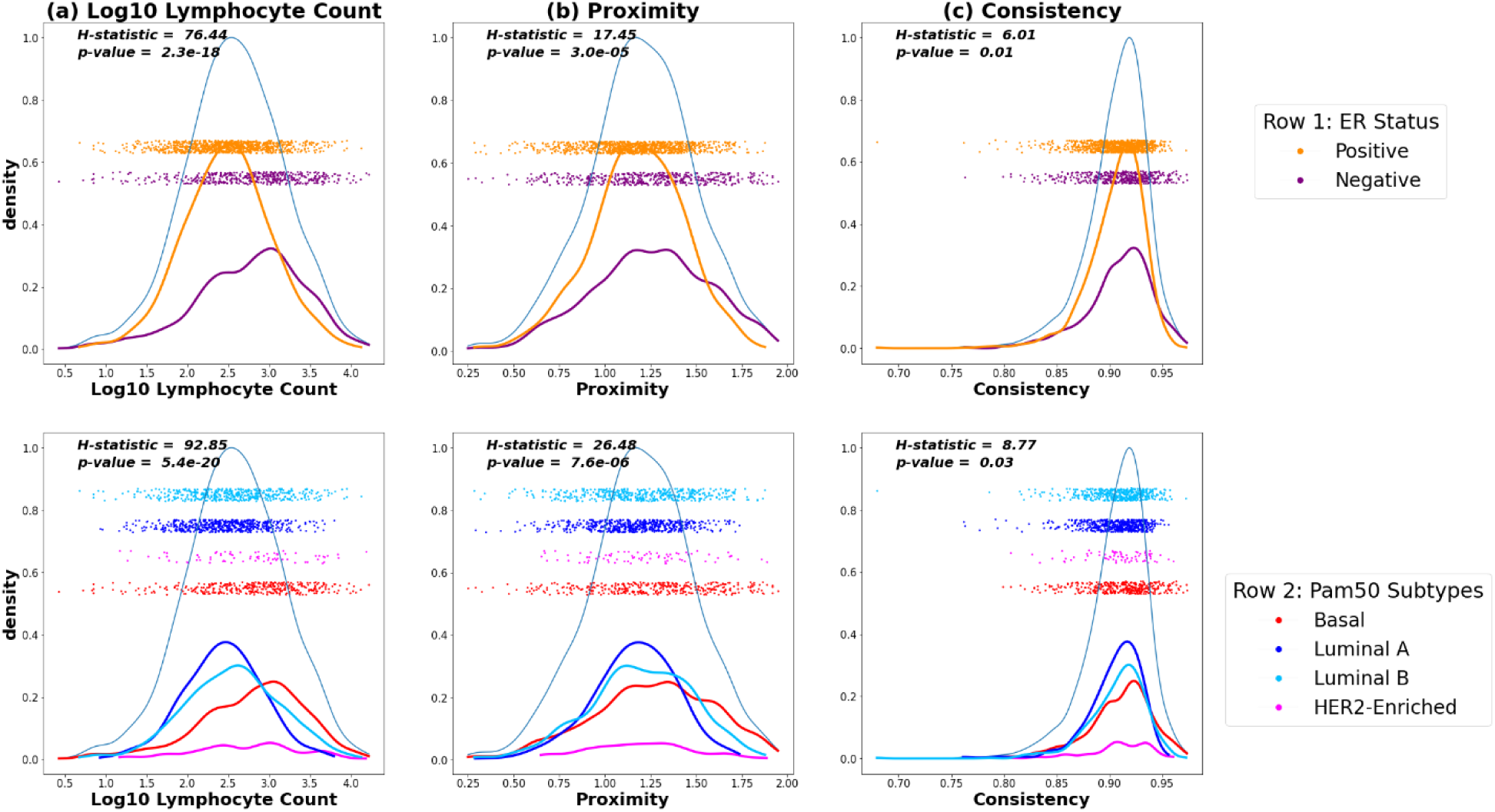
Density plots for the (a) log base 10 lymphocyte counts, (b) proximity, and (c) consistency values in Phase 3 CBCS participants with sub-densities and overlayed with jittered scatter plot of the data colored by ER status (top row) and PAM50 subtypes (bottom row). H statistics and p-values correspond to the Kruskal-Wallis test, a non-parametric version of ANOVA.

We also determined if there were statistically significant differences between the median of the count, proximity, and consistency by PAM50 molecular subtypes (Figure 2). Similarly, the difference between the median of the log10 lymphocyte count among PAM50 subtypes (H = 92.85, p-value = 5.4e-20) was more significant than that of proximity and consistency (H = 26.48, p-value = 7.6e-6 and H = 8.77, p-value = 0.03 respectively). As with ER status, the distributions of proximity and consistency were overlapping, notably in Basal vs. Luminal. This implies that while proximity and consistency does not distinguish ER status and PAM50 subtypes as well as lymphocyte count, they identify spatial parameters that are shared across subtypes. This was also true when including ‘double positive’ (CD8+ and FoxP3+) cells to compute proximity and consistency (Supplementary Figure 1).

### Recurrence Free Survival Analysis

Participants were stratified into high and low categories for proximity, consistency, and lymphocyte count based on first splitting into four groups at the quartiles and collapsing groups with similar hazard ratios. Kaplan-Meier estimates of the Recurrence-Free Survival (RFS) functions are shown in Figure 3, with hazard ratios for the univariate and multivariate Cox proportional hazards model in Figure 4.

**Figure 3:**
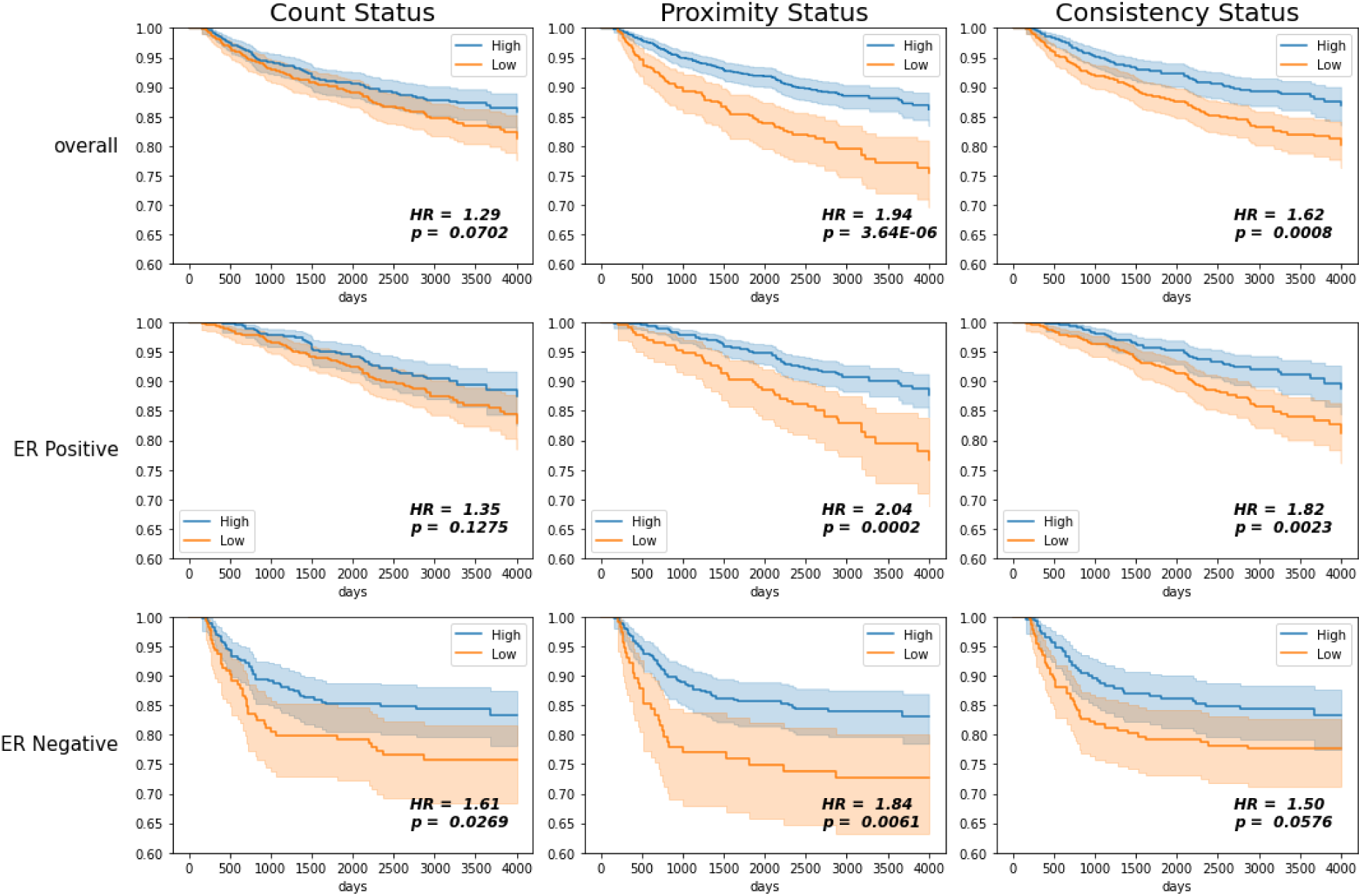
Kaplan-Meier estimates for recurrence-free survival (RFS) comparing high vs. low proximity, consistency, and lymphocyte count in Phase 3 CBCS participants overall and stratified by ER status. 95% confidence bounds are given by the shaded region. P-values correspond to the log-rank test of the difference between curves. Hazard Ratios (HR) are given for the difference between the hazards of the survival curves of the high vs. low status.

**Figure 4:**
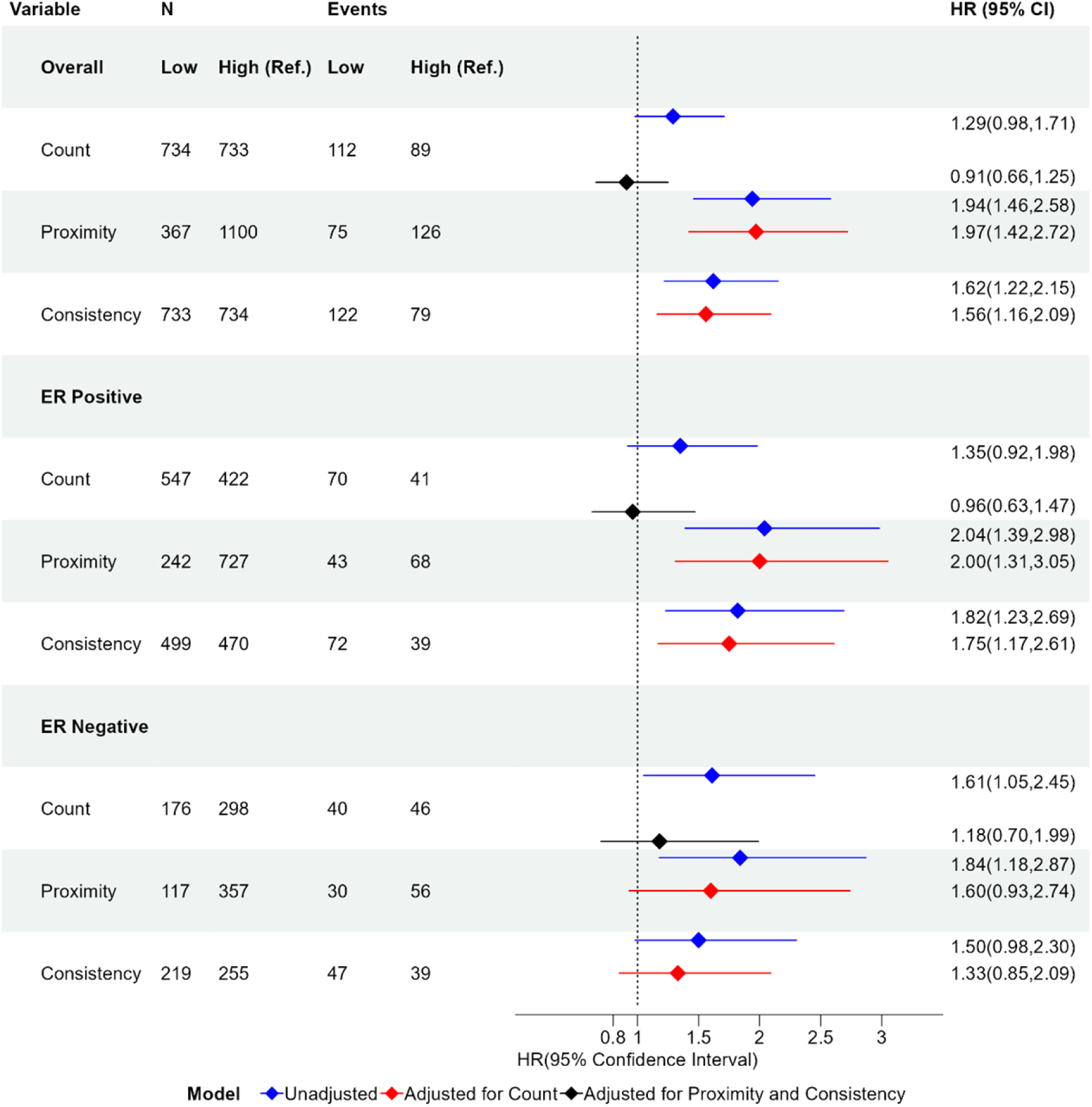
Forest plot of hazard ratios and 95% confidence intervals for proximity, consistency, and lymphocyte count binary variables comparing low and high values when considering all participants and stratified by ER status. The Hazard ratio is given in the unadjusted case (blue), adjusted for lymphocyte count (red), and adjusted for both proximity and consistency (black). The hazard ratio is with respect to the reference group (Ref.). The references groups are High Count, High Proximity, and High Consistency. Group size (N) and number of participants who experience recurrence (Event) are given for each group. HR: Hazard Ratio; 95% CI: 95% confidence Interval.

Consistent with a priori knowledge, ER negative tumors showed associations between lymphocyte counts and improved outcomes (Figure 3, third row, p=0.027). Higher lymphocyte count was associated with longer survivorship, with low counts showing a hazard ratio of 1.61 (95% CI [1.05, 2.45] vs. High counts). However, proximity also showed associations, with HRs that were slightly stronger (1.84, 95% CI [1.18, 2.87]). Consistency was only borderline significantly associated with recurrence (p-value of 0.0576).

Associations diverged further when considering ER+ and ER-tumors together. Among breast cancers overall, high proximity and high consistency were both significantly associated with a longer time to recurrence (p-values < 0.001), while lymphocyte count was not significantly associated with recurrence (Figure 3, top row). Low proximity participants had a recurrence rate nearly twice that of high proximity participants, with a hazard ratio of 1.94 (95% CI [1.46, 2.58]). Low consistency participants had a higher chance of recurrence, with a hazard ratio of 1.62 (95% CI [1.22, 2.15] vs. high consistency). Lymphocyte count had the weakest association overall, with a hazard ratio of 1.29 (95% CI [0.98, 1.71]) that was not statistically significant (p-value = 0.07).

Among ER positive participants, high proximity and consistency were also significantly associated with longer time to recurrence (Figure 3, middle row, p-values < 0.01). The hazard ratios for low vs high proximity and consistency are 2.04 (95% CI [1.39, 2.98]) and 1.82 (95% CI [1.23, 2.69]) respectively, and therefore were similar to those for cancers overall. In contrast, the hazard ratio for low vs high lymphocyte count was 1.35 (95% CI [0.92, 1.98]) and was not statistically significant (Figure 4). This suggests that while lymphocyte count is not associated with recurrence in ER positive participants, spatial characteristics as described by proximity and consistency are consistently associated with recurrence irrespective of ER status.

We next sought to assess the independent value of proximity and consistency relative to models with lymphocyte count alone. We used nested Cox Proportional Hazards models and Likelihood Ratio Tests (LRT) to determine whether proximity and consistency remained significant in a model with lymphocyte count (Supplemental Table 1). We found that the model with count, proximity, and consistency had a significantly higher likelihood than the model with count alone (p-value =1.57e-05). Furthermore, when adjusting for count, the hazard ratios for proximity and consistency did not differ significantly from the unadjusted hazard ratios in both overall and ER positive cohorts (Figure 4).

Similarly, we assessed whether lymphocyte count provided independent value in models that include proximity and consistency. Compared to a model that included proximity and consistency only, LRT tests showed that the full model (with count) did not differ statistically from the reduced model (with proximity and consistency only, p-value = 0.55). Moreover, when adjusting the effects of count by including proximity and consistency, the hazard ratio decreased significantly (Figure 4). Even among ER negatives, the hazard ratio for count was no longer significant after adjusting for proximity and consistency (1.18, 95% CI [0.70,1.99]). This suggests that, even in ER negative participants, spatial characteristics of TILs may provide information beyond what can be inferred strictly from the number of lymphocytes.

The relationship between count and proximity does show a correlation, and as a result we also considered stratified models to assess the relationship between count and proximity (Figure 5). Among participants with low lymphocyte counts, high proximity was associated with improved outcomes (p-value = 0.0002). Among participants with high lymphocyte counts, high consistency remained associated with improved survival (p-value of 0.0005). Thus, proximity and consistency have independent value beyond lymphocyte count in predicting recurrence.

**Figure 5:**
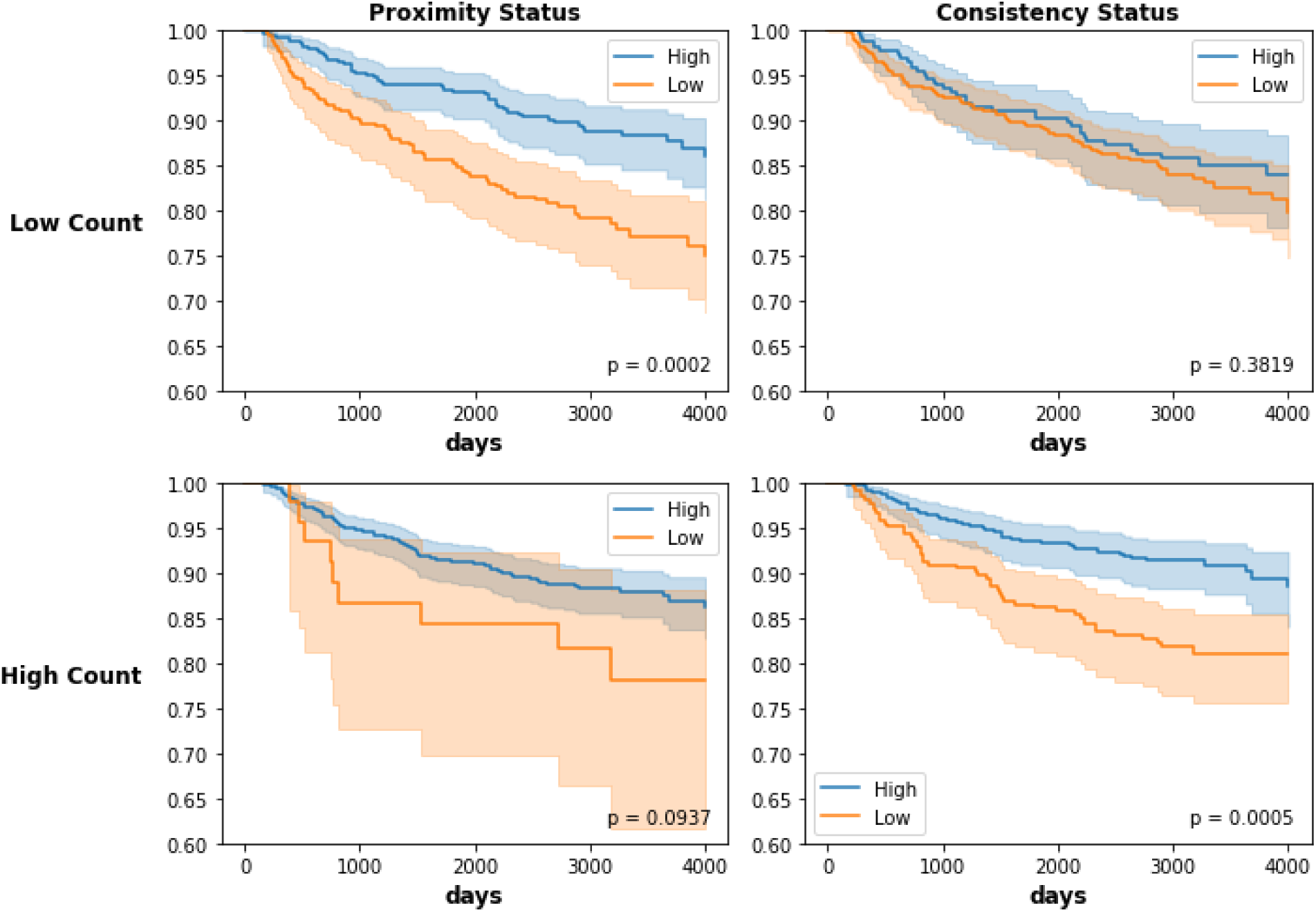
Kaplan-Meier estimates for recurrence-free survival comparing high vs. low proximity and consistency in Phase 3 CBCS participants overall and stratified by high vs low lymphocyte count. 95% confidence bounds are given by the shaded region. P-values correspond to the log-rank test of the difference between curves. The difference in RFS curves for high vs. low proximity is more significant in participants with low lymphocyte count. The difference in RFS curves for high vs. low consistency is more significant in participants with high lymphocyte count.

### Comparison of TMA based spatial assessment of TILs to Whole Slide TIL analyses

To contextualize our measures of immune spatial relationships relative to established metrics, we compared the proximity and consistency measurements (determined based on TMAs from the Carolina Breast Cancer Study) to a set of TIL scores from the slides of the same patients. In the slides, larger scale metrics including TILs forests, TILs deserts, intratumoral strength, and peritumoral strength were estimated as described by Fassler.^20^ It is notable that proximity and consistency can be determined in 1mm diameter TMA cores, whereas the whole slide images (WSI) evaluated for forests and deserts consist of tissue samples which are 15-25mm across the longest dimension. On this larger scale, TIL forest and desert scores address presence or absence of TILS, respectively and intratumoral strength and peritumoral strength describe the level of TILs in the tumor interior and periphery, respectively.

Proximity, consistency, and lymphocyte count were all significantly correlated with the four WSI TIL scores (Figure 6). We observed the strongest correlations with intratumoral strength (corr. = 0.598), but also observed associations with pertumoral strength (corr. = 0.567) and TIL forests (corr. = 0.503) and an inverse association with TIL deserts (corr. = −0.469) (Figure 6). Consistency had a weaker correlation due to a parabolic relation with intratumoral and peritumoral strength; that is, when lymphocytes are absent or abundant, consistency is high and variation is observed predominantly among sparsely distributed lymphocytes. As proximity scores increase beyond 1, consistency increases. Still, consistency was correlated with TIL forests (corr. = 0.321) and inversely with TIL deserts (corr. = −0.357) scores, but more weakly than proximity and lymphocyte count. Taken together, these analyses suggest that our small-scale spatial metrics from TMAs reflect prognostically relevant, large-scale TIL spatial structures.

**Figure 6:**
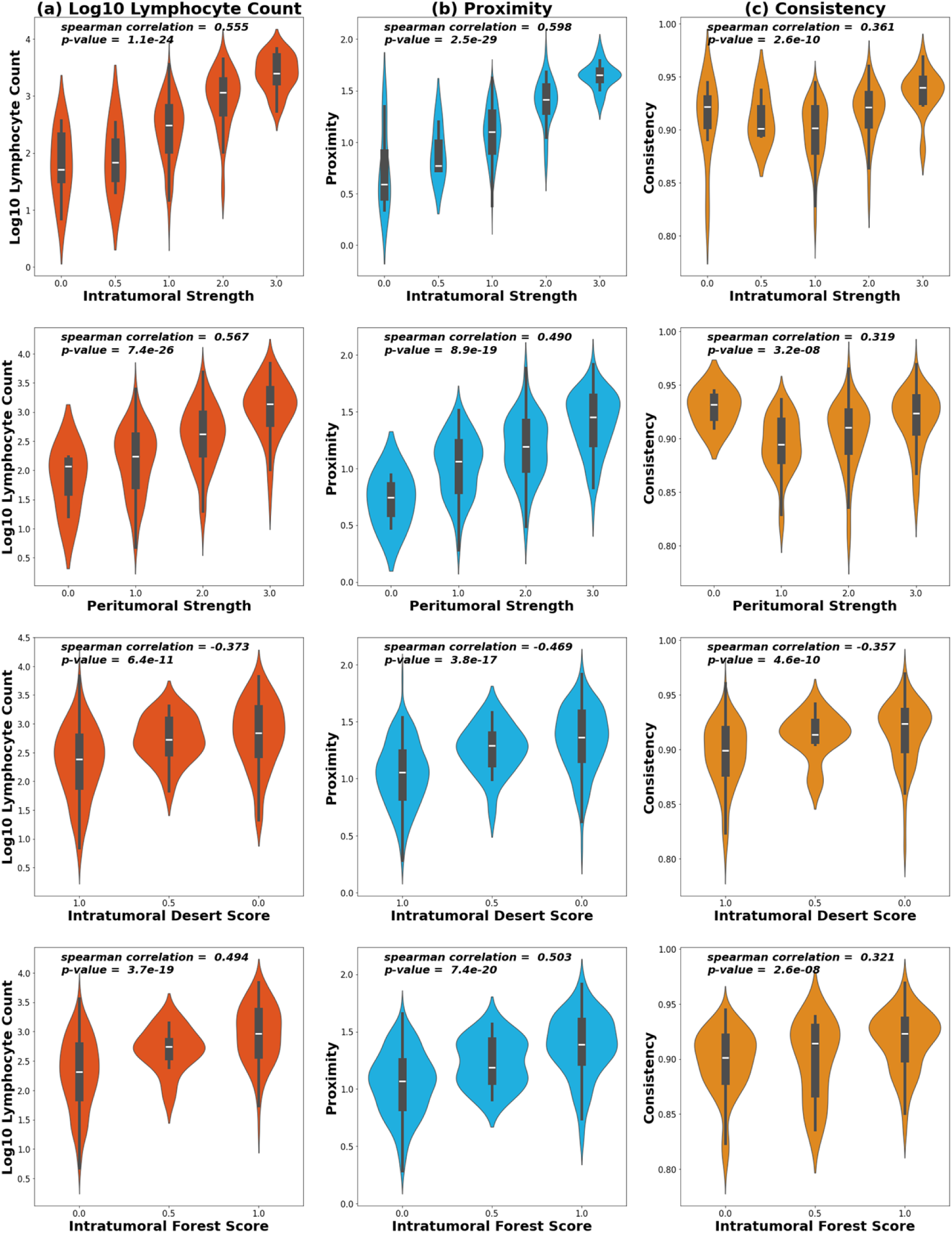
Violin plots depicting the distribution of (a) proximity, (b) consistency, and (c) log10 count for the scores of, intratumoral strength, and peritumoral strength, TIL Deserts, and TIL Forests. Spearman’s correlation coefficient and p-values for coefficient significance are given. Note that only 288 of the total participants were scored for TIL Forests, Deserts, intratumoral strength, and peritumoral strength.

The WSI TIL scores, proximity, and consistency were all included as covariates in a cox proportional hazards model. A smaller sample size when including the WSI scores results in univariate hazard ratios that are different from those previously listed. The results are included in supplemental Table 4. The hazard ratios for proximity and consistency after adjusting for WSI TIL scores are 2.31 (95% CI [1.25, 4.29]) and 1.57 (95% CI [0.91, 2.71]) respectively, compared to the unadjusted hazard ratios of 1.94 (95% CI [1.46, 2.58]) for proximity and 1.62 (95% CI [1.22, 2.15]) for consistency. Though the hazard ratio for consistency is no longer significant, proximity remains significant after adjusting for WSI TIL scores. Moreover, the only statistically significant covariate in the model is proximity (p-value of 0.01). Therefore, proximity provides the most significant information for determining RFS even when including all WSI TIL scores.

### Relation to Gene Expression Biomarkers

We next hypothesized that spatial parameters for immune infiltration may predict molecular features as measured in RNA. We queried both immune signatures and signatures of genomic instability. Hamilton et al. 2022 identified three distinct immune classes (Adaptive-Enriched, Innate-Enriched, or Immune Quiet) based on gene expression that were associated with TIL counts^19^. As shown in Table 1, Adaptive-Enriched (AE) tumors were associated with high proximity, consistency and lymphocyte count compared to the combined Innate-Enriched (IE) and Immune Quiet (referent) with odds ratios of 4.9 (95% CI [3.2,7.7]), 2.4 (95% CI [1.8,3.2]), and odds ratio of 6.0 (95% CI [4.4,8.4]) respectively. Additionally, a biomarker of Any Genomic Instability (AGI) biomarker (defined by high homologous recombination deficiency (HRD) and/or TP53 Mutant-like signatures; vs. No Genomic Instability markers), was associated with Adaptive immune class in previous work^12^, and was associated with high lymphocyte count [OR =2.6 (95% CI [1.9, 3.3])] but less strongly associated with high proximity and consistency, with odds ratios that were not significant at the 95% confidence level.

**Table 1:**
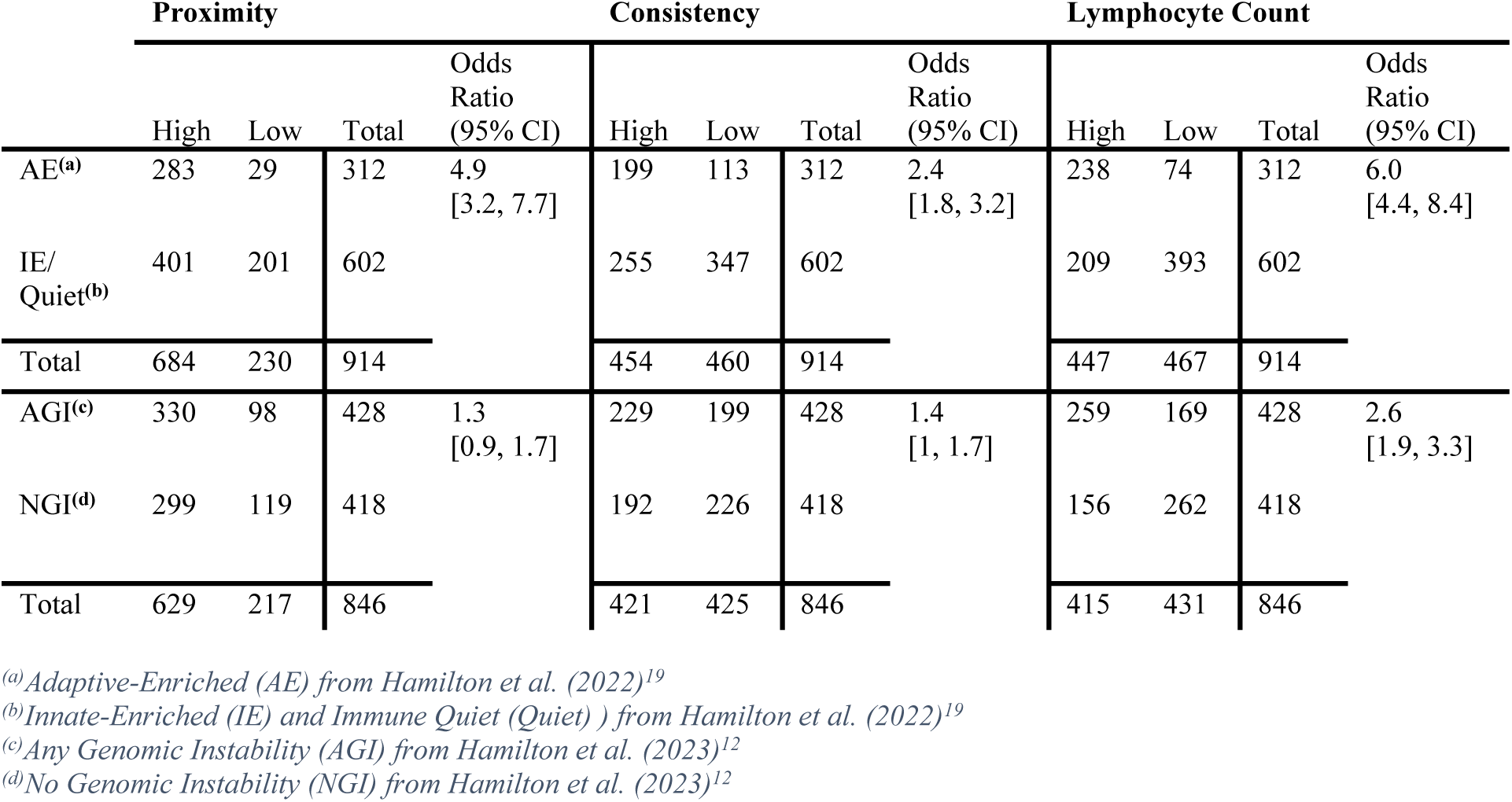
Contingency Tables for Spatial Biomarkers vs Gene Expression Derived Immune Classes.

We also considered the immune classes along with proximity and consistency in a cox proportional hazards model (supplemental Table 5). After adjusting for the immune classes, the hazard ratios for proximity and consistency are 1.96 (95% CI [1.31, 2.93]) and 1.45 (95% CI [1.00, 2.10]) respectively. These hazard ratios are relatively unchanged compared to the univariate hazard ratios of 1.94 (95% CI [1.46, 2.58]) for proximity and 1.62 (95% CI [1.22, 2.15]) for consistency. Furthermore, the most significant covariates are proximity (p-value < 0.005) and consistency (p-value of 0.05).

## Discussion

While the clinical significance of immune response to breast tumors is established, there is remaining uncertainty about the importance in non-triple negative subtypes (i.e. ER-positives) and the best metrics for identifying immune susceptibility are still debated. Our work explored spatial parameters of CD8+/FOXP3-cells in tumor and peritumoral regions as predictors of recurrence and in association with gene expression signatures. Proximity and Consistency were significant determinants of recurrence for both ER+ and ER-breast cancers, while lymphocyte count showed weaker associations, especially among ER+, and did not offer independent value in models with proximity and consistency. When inflammation is high, there may be little prognostic distinction between lymphocyte counts and proximity; that is, when lymphocyte count is very high, proximity is also high. In these cases, Consistency or uniformity of TILs may be a better gauge of an effective immune response when count is high. Thus, among participants with high counts, we observed that high consistency was associated with improved recurrence-free survival, whereas low proximity was rare and not associated with recurrence. However, in cases where lymphocyte count was low, high spatial proximity was important and associated with improved recurrence-free survival. These data suggest that a lower level, but well-targeted immune response, can have important efficacy in responding to tumors. Furthermore, proximity and consistency were both significantly correlated with other spatial TILs scores based on whole slide images and with Adaptive-Enriched gene expression. These results imply that the location of T-cells in relation to tumor cells have prognostic and biological significance beyond bulk lymphocytic infiltration and underscores the importance of spatial parameters in characterization of TIME.

The role of CD8+ T-cells in the TIME is well established. Several previous studies demonstrated that higher levels of CD8+ T-cells are associated with positive prognosis in TNBC.^24,25^ However, most of these studies have suggested limited benefit of CD8+ infiltration for ER+ participants^26,27^. One study documented CD8 T-cells in both stroma and intratumoral regions and established an association with survival for both ER+ and ER-participants.^28^ Another study used QuPath, an open-source digital pathology software, to build a machine learning algorithm for scoring H&E images based on density of TILs in various tissue compartments^29^ and confirmed prognostic significance of TILs in TNBC, but did not consider ER+ or ER-/HER2+ breast cancer.^29^ None of these studies focused on spatial proximity and consistency, instead emphasizing CD8+ T-cell counts. Inconsistencies in associations with ER+ disease in the previous literature may reflect unmeasured parameters; our data suggests that proximity and consistency are two plausible parameters. For CD8+ T cells to induce apoptosis, i.e. cell death, they must be in direct contact with tumor cells^16^, and therefore distance-based methods have theoretical advantages for naturally quantifying interactions between CD8+ T cells and tumor cells.

Methods of automatic spatial characterization of TILs have been developed in various studies for the analysis of H&E images.^20,29–31^ Abousamra et al. used convolutional neural networks to create whole slide image TILs maps by classifying tiled image patches as TIL positive or negative (based on at least 2 lymphocytes detected).^31^ These models have advantages in using complex deep learning models.

However, classifying image tiles could miss fine-grained variation in the TIME and lead to misclassification of proximity. Our use of simple distance-based methods eases scalability to whole slide images and is well-suited to TMAs that are commonly available for large cohorts. Moreover, proximity and consistency of lymphocyte infiltration at the TMA level is significantly correlated with measures of TIL intratumoral strength and peritumoral strength at the whole-slide level. This suggests that spatial metrics applied to TMA are predictive of large-scale spatial immune structures within the tumor.

This work has some noteworthy strengths and limitations. A strength of our analysis was the ability to compare the spatial metrics from IF images to bulk gene expression data from the same specimens. The connection between gene expression derived immune clusters^19^ and the spatial biomarkers suggests that CD8 T-cells function in the TIME reflect broader patterns of immune cells. However, we were unable to fully interrogate the relationship between multiple immune markers and spatial arrangement. Proximity and consistency could be affected by interactions with other lymphocyte types, with some previous work suggesting that increased regulatory T-cells could impair CD8 T-cell activity. Furthermore, we observed that genomic instability based on TP53 and HRD^12^ did not predict proximity and consistency of immune cells as strongly as count. Genomic instability-associated neo-antigen production may be required for a robust immune response. However, we lack data to specifically interrogate neoantigens as a mediator of count vs. spatial immunogenic patterns. Finally, while IF allowed us to distinguish lymphocyte types, we studied CD8+ cells and neglected other spatial interaction between immune cell types. However, CD8 is a priority marker given its established role in cytotoxic immune responses.

In summary, our analysis suggests prognostic value in the location of lymphocytes, specifically CD8+ T-cells. If validated, proximity and consistency may identify ER positive breast cancers that benefit from immune therapy, expanding treatment options for as many as 75% of ER+ tumors (i.e. the proportion in this dataset that showed high proximity). Given that ER+ tumors represent the majority of breast tumors, closer evaluation of immune response to identify possible beneficiaries is highly impactful.

## Data Availability

CBCS data are available upon request (https://unclineberger.org/cbcs).

## Acknowledgements

This research was supported by a grant from UNC Lineberger Comprehensive Cancer Center, which is funded by the University Cancer Research Fund of North Carolina, the Susan G Komen Foundation (OGUNC1202, OG22873776, SAC210102, TREND21686258), National Cancer Institute (R01CA253450), the National Cancer Institute Specialized Program of Research Excellence (SPORE) in Breast Cancer (NIH/NCI P50-CA058223), the Breast Cancer Research Foundation (HEI-23-003), and the US Department of Defense (HT94252310235). This research recruited participants &/or obtained data with the assistance of Rapid Case Ascertainment, a collaboration between the North Carolina Central Cancer Registry and UNC Lineberger. RCA is supported by a grant from the National Cancer Institute of the National Institutes of Health (P30CA016086). The findings and conclusions in this publication are those of the authors and do not necessarily represent the views of the North Carolina Department of Health and Human Services, Division of Public Health. The authors would like to acknowledge the University of North Carolina BioSpecimen Processing Facility for sample processing, storage, and sample disbursements (http://bsp.web.unc.edu/) and the Breast Cancer Research Foundation HEI-23-003. We are grateful to CBCS participants and study staff.

## Statement of Ethics

The study was approved by the University of North Carolina Institutional Review Board in accordance with U.S. Common Rule. All study participants provided written informed consent prior to study entry. This study complied with relevant ethical regulations, including the Declaration of Helsinki.

## Supplemental Materials

**Figure 1:**
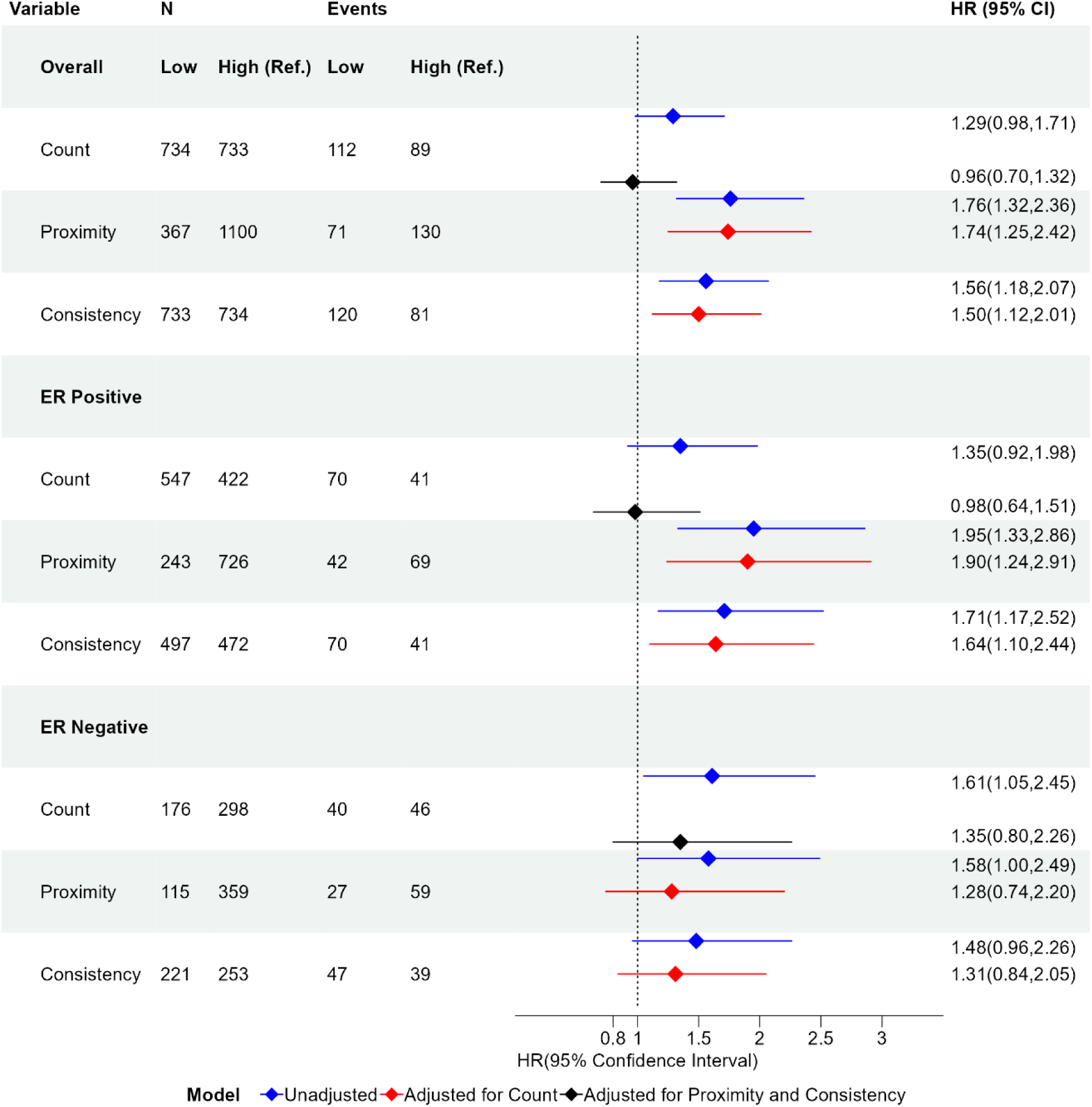
Results when using CD8 only and CD8 and FoxP3 double positive cells to compute proximity and consistency. Forest plot of hazard ratios and 95% confidence intervals for proximity, consistency, and lymphocyte count binary variables comparing low and high values when considering all participants and stratified by ER status. The Hazard ratio is given in the unadjusted case, adjusted for lymphocyte count, and adjusted for both proximity and consistency. The hazard ratio is with respect to the reference group (Ref.). The references groups are High Count, High Proximity, and High Consistency. Group size (N) and number of participants who experience recurrence (Event) are given for each group. HR: Hazard Ratio; 95% CI: 95% confidence Interval.

**Table 1:** Summary of multivariate cox proportional hazards models with proximity, consistency, and lymphocyte count binary covariates for all participants. Estimates for the covariate adjusted hazard ratios are provided as well as p-values for the significance of the hazard ratios. Models with nested covariates are compared using the likelihood ratio test and p-values for the significance of the complex model compared to the nested model are provided. The reference group for each of the covariates are high proximity, high consistency, and high lymphocyte count respectively.

| Models | Covariates | Estimated Hazard Ratio | Estimate p-value | p-value for Likelihood Ratio Test |  |  |
| --- | --- | --- | --- | --- | --- | --- |
|  |  |  |  | Model 3 | Model 4 | Model 5 |
| 1 | Low proximity | 1.94 | 5.43e-06 | 0.016 | 0.88 |  |
| 2 | Low lymphocyte count | 1.29 | 0.071 |  | 6.33e-05 | 1.57e-05 |
| 3 | Low proximity | 1.77 | 0.00015 |  |  | 0.55 |
|  | Low consistency | 1.43 | 0.017 |  |  |  |
| 4 | Low lymphocyte count | 0.98 | 0.88 |  |  | 0.013 |
|  | Low proximity | 1.96 | 5.11e-05 |  |  |  |
| 5 | Low proximity | 1.85 | 0.00025 |  |  |  |
|  | Low consistency | 1.45 | 0.014 |  |  |  |
|  | Low lymphocyte count | 0.91 | 0.55 |  |  |  |

**Table 2:** The tuning parameters used for cell detection and thresholds for cell classification in the QuPath functions.

| QuPath Cell detection Tuning Parameters |  |
| --- | --- |
| Channel Used for Detection | Hoechst |
| Requested Pixel Size | 0.5 |
| Background Radius | 8 |
| Median Filter Radius | 0 |
| Sigma | 1.5 |
| Minimum Area | 10 |
| Maximum Area | 400 |
| Threshold | 11 |
| Cell Expansion | 3 |
| QuPath Classification Thresholds |  |
| CD8 Positive Threshold | 25.12 |
| FoxP3 Positive Threshold | 80 |
| CK Positive Threshold | 31.62 |

**Table 3:** Results from attempt to find optimal cutpoint for count, proximity, and consistency using a 2/3 to 1/3 training and test split in the overall dataset, the ER positive only, and the ER negative only. Hazard Ratios (HR) and p-values for the log-rank test are given for all cases in both the training and testing data. Results for testing data is based on the optimal cutpoint found using the training set.

|  |  | Training |  | Testing |  |
| --- | --- | --- | --- | --- | --- |
|  | High (ref) vs. Low | HR | p-value | HR | p-value |
| **overall** | Count | 2.07 | 0.0001 | 1.65 | 0.0792 |
|  | Proximity | 1.9 | 0.0003 | 2.02 | 0.0044 |
|  | Consistency | 1.69 | 0.0101 | 1.06 | 0.8503 |
| **ER Positive** | Count | 3.19 | 3.35E-07 | 1.54 | 0.2393 |
|  | Proximity | 2.58 | 0.0001 | 1.87 | 0.0709 |
|  | Consistency | 2.3 | 0.0002 | 0.87 | 0.7256 |
| **ER Negative** | Count | 5.9 | 2.84E-06 | 1.46 | 0.307 |
|  | Proximity | 7.59 | 0.0009 | 2 | 0.1888 |
|  | Consistency | 5.63 | 0.001 | 2.8 | 0.0775 |

The above table shows the results from optimizing the cutpoint for the high vs. low count, proximity, and consistency. We split the data based on a 2/3 to 1/3 training and testing split. Furthermore, we used stratified sampling on the recurrence data to ensure there would be an adequate number of participants with recurrence in the training and testing sets. We find an optimal cutpoint based on the quantile of the data which minimizes the p-value for the log-rank test. Though the cutpoints appear to perform well on the training set, in most cases there appears to be overfitting based on very different hazard ratios in the testing data. Furthermore, many results which were statistically significant in the training data are no longer statistically significant in the testing data.

**Table 4:**
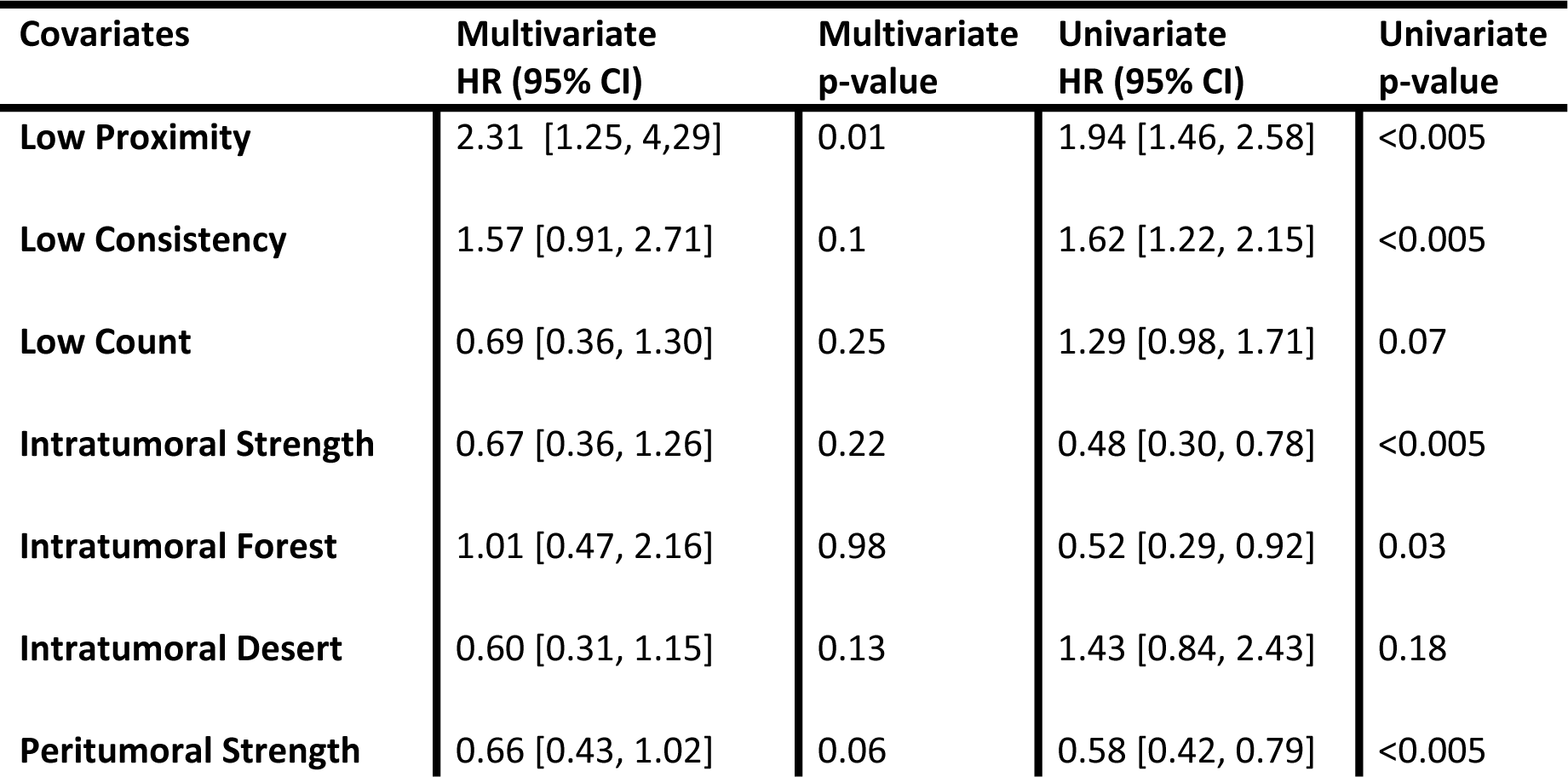
Cox Proportional Hazards Model with WSI TIL Scores.

| Covariates | Multivariate<br>HR (95% CI) | Multivariate<br>p-value | Univariate<br>HR (95% CI) | Univariate<br>p-value |
| --- | --- | --- | --- | --- |
| Low Proximity | 2.31 [1.25, 4.29] | 0.01 | 1.94 [1.46, 2.58] | <0.005 |
| Low Consistency | 1.57 [0.91, 2.71] | 0.1 | 1.62 [1.22, 2.15] | <0.005 |
| Low Count | 0.69 [0.36, 1.30] | 0.25 | 1.29 [0.98, 1.71] | 0.07 |
| Intratumoral Strength | 0.67 [0.36, 1.26] | 0.22 | 0.48 [0.30, 0.78] | <0.005 |
| Intratumoral Forest | 1.01 [0.47, 2.16] | 0.98 | 0.52 [0.29, 0.92] | 0.03 |
| Intratumoral Desert | 0.60 [0.31, 1.15] | 0.13 | 1.43 [0.84, 2.43] | 0.18 |
| Peritumoral Strength | 0.66 [0.43, 1.02] | 0.06 | 0.58 [0.42, 0.79] | <0.005 |

**Table 5:** Cox Proportional Hazards Model with Gene Expression based Immune Classes.

| Covariates | Multivariate<br>HR (95% CI) | Multivariate<br>p-value | Univariate<br>HR (95% CI) | Univariate<br>p-value |
| --- | --- | --- | --- | --- |
| Low Proximity | 1.88 [1.25, 2.84] | <0.005 | 1.94 [1.46, 2.58] | <0.005 |
| Low Consistency | 1.42 [0.98, 2.06] | 0.06 | 1.62 [1.22, 2.15] | <0.005 |
| Low Count | 0.81 [0.53, 1.22] | 0.31 | 1.29 [0.98, 1.71] | 0.07 |
| CD8 T-Cell Signature | 0.92 [0.78, 1.07] | 0.28 | 0.84 [0.73, 0.96] | 0.01 |

**Table 6:**
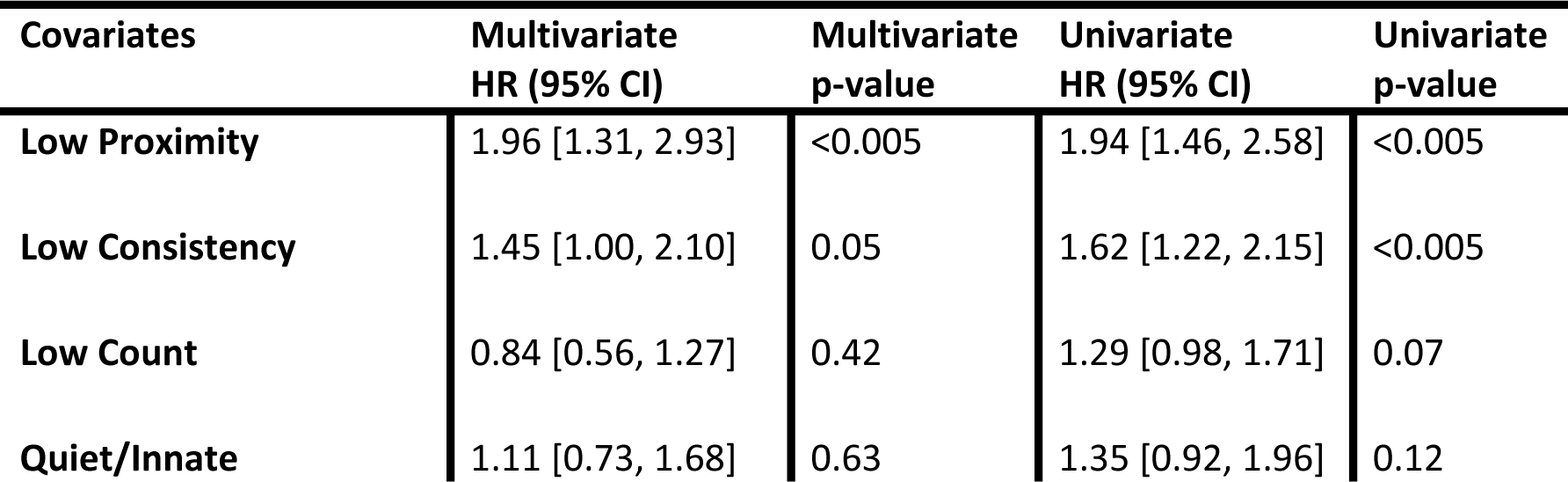
Cox Proportional Hazards Model with CD8 T-cell Signature.

| Covariates | Multivariate<br>HR (95% CI) | Multivariate<br>p-value | Univariate<br>HR (95% CI) | Univariate<br>p-value |
| --- | --- | --- | --- | --- |
| Low Proximity | 1.96 [1.31, 2.93] | <0.005 | 1.94 [1.46, 2.58] | <0.005 |
| Low Consistency | 1.45 [1.00, 2.10] | 0.05 | 1.62 [1.22, 2.15] | <0.005 |
| Low Count | 0.84 [0.56, 1.27] | 0.42 | 1.29 [0.98, 1.71] | 0.07 |
| Quiet/Innate | 1.11 [0.73, 1.68] | 0.63 | 1.35 [0.92, 1.96] | 0.12 |

